# Quantitative faecal immunochemical test for patients with ‘high risk’ bowel symptoms: a prospective cohort study

**DOI:** 10.1101/2020.05.10.20096941

**Authors:** Helga E Laszlo, Edward Seward, Ruth M Ayling, Jenny Lake, Aman Malhi, Allan Hackshaw, Clare Stephens, Kathy Pritchard-Jones, Donna Chung, Michael Machesney

## Abstract

**Objectives:** To evaluate whether quantitative measurement of faecal haemoglobin (f-Hb) using faecal immunochemical testing (FIT) can be used to rule out colorectal cancer (CRC) for patients who present to primary care with ‘high risk’ symptoms defined by national guidelines for urgent referral for suspected cancer (NICE NG12).

**Design:** Prospective cohort study carried out between April 2017 and March 2019.

**Setting:** 59 GP practices in London and 24 hospitals in England.

**Participants:** Symptomatic patients in England referred to the urgent CRC pathway who provided a faecal sample for FIT in addition to standard investigations for cancer.

**Main outcome measures:** CRC was confirmed by established clinical and histopathology procedures. f-Hb (μg per gram of stool) was measured in a central laboratory blinded to cancer outcome. We calculated sensitivity (percentage of patients with CRC who have f-Hb exceeding specified cut-offs); false-positive rate [FPR] (percentage of patients without CRC whose f-Hb exceeds the same cut-offs); and positive predictive value [PPV] (percentage of all patients with f-Hb above the cut-offs who have CRC).

**Results:** 4676 patients were recruited of whom 3596 patients were included (had a valid FIT test and a known definitive diagnosis). Among the 3596, median age was 67 years, 53% were female and 78% had colonoscopy. 90 patients were diagnosed with CRC, 7 with other cancers, and 3499 with no cancer found. f-Hb did not correlate with age, sex or ethnicity. Using f-Hb ≥4μg/g (lowest limit of detection), sensitivity, FPR and PPV were 87.8%, 27.0% and 7.7% respectively. Using f-Hb ≥10μg/g, the corresponding measures were 83.3%, 19.9% and 9.7%. 15 patients with CRC had f-Hb below 10μg/g. If FIT had been used at thresholds of 10μg/g or 4μg/g, 1 in 6 or 1 in 8 patients with cancer respectively would have been missed. If the absence of anaemia or abdominal pain is used alongside f-Hb 10 μg/g, only 1 in 18 cancers would be missed but 56% of people without CRC could potentially avoid further investigations including colonoscopies.

**Conclusions:** In our study, if FIT alone had been used to determine urgent referral for patients with ‘high risk’ symptoms for definitive cancer investigation, some patients with bowel cancer would not have been diagnosed. If used in conjunction with clinical features, particularly in the absence of anaemia, the efficacy of FIT is significantly improved. With appropriate safety netting, FIT could be used to focus secondary care diagnostic capacity on patients most at risk of CRC.

## INTRODUCTION

Colorectal cancer (CRC) is the fourth most common cancer in the UK, and the second largest cause of death due to cancer. If diagnosed at an early stage, long term survival is over 90%.^1^ Typically, individuals who see a primary care physician because of high risk lower abdominal symptoms, are referred for investigations for lower gastrointestinal cancer. In the UK for example, these symptoms are defined in the National Institute of Health and Care Excellence (NICE) diagnostic guideline (NG12), used for referral to an urgent referral pathway (also known as “2-week wait”) to be seen by a specialist within 14 days.^2^ In 2018/19 over 396,000 patients in England were seen by a specialist as part of this pathway.^3^ The majority of these patients were investigated with colonoscopy, the gold standard for detecting colorectal disease including CRC, higher-risk adenoma (HRA), inflammatory bowel disease (IBD) and other benign conditions. However, less than 8% of patients with high risk symptoms have CRC.^4^

Several studies indicate that faecal haemoglobin (f-Hb) has a higher predictive value than symptoms of colorectal disease.^5–7^ Moreover, the urgent referral pathway has not improved survival rates compared to other pathways for patients with CRC^4,8^ whilst detection through the national bowel cancer screening programme is associated with higher survival rates.^9^ There is a need for an effective triage test embedded in primary care that provides rapid and simple assessment of symptomatic patients. This may facilitate early diagnosis of CRC and improve cancer survival whilst reducing the number of unnecessary diagnostic investigations on people with symptoms but who do not have CRC.^10^

Several studies have examined the faecal immunochemical test (FIT), which quantifies faecal haemoglobin (f-Hb) concentration, and its ability to reliably exclude CRC, HRA and IBD in asymptomatic (i.e. screening) populations^11–13^, and symptomatic ‘lower risk’^14–17^ and ‘higher’ risk^10,18–22^ patient groups. NICE guidance currently recommends the use of FIT to triage ‘low’ risk patients (those who do not meet the NG12 urgent pathway referral criteria) presenting with lower abdominal symptoms in primary care^23^, where a f-Hb concentration of ≥10μg/g can be used to justify an urgent referral.

A meta-analysis of FIT studies indicated that f-Hb has a higher test performance than the NG12 referral pathway for all significant colorectal disease.^7^ Westwood et al (2017) found that referral directly to colonoscopy is less cost-effective than using FIT to triage the use of colonoscopy.^24^

Three studies conducted in a UK population suggested that a ‘negative’ FIT test (undetectable f-Hb or concentration <10μg/g) could ‘rule out’ colorectal cancer^18–20^ in symptomatic patients referred for cancer investigations, with a significant reduction in the number of unnecessary colonoscopies. Two reported that 100% of CRC cases could be detected and one had a sensitivity of 83%, with specificities ranging between 43 and 93%. However, all of these studies were relatively small (430 to 755 patients), each with only 11 to 28 CRC cases and consequently imprecise estimates of sensitivity. Studies conducted in other countries have the same limitations. Therefore, it remains inconclusive whether using FIT alone leads to an unacceptable number of missed cancers in symptomatic patients.

Factors such as younger age^25–27^, female gender^26–28^, anaemia^10,20^, and degree of deprivation^29^ might be associated with lower f-Hb among asymptomatic populations which could lead to higher false negative FIT results. There is limited evidence on whether these factors contribute to more false negative FIT tests among symptomatic patients, and NICE has highlighted the need for further research.^23^

We evaluated the test performance of FIT among a large cohort of patients referred from primary care with lower gastrointestinal symptoms on the NG12 urgent referral pathway.

## METHODS

### Study design

We conducted a prospective multi-centre observational study (the qFIT study), which recruited patients from 24 hospitals in England and 59 general practices in London between April 2017 and March 2019. Primary and secondary care sites were invited through the National Institute for Health Research Clinical Research Network (NIHR CRN). The participating sites were selected to cover a wide geography within and outside of London to ensure diversity (see Supplementary Table A for the full list). National ethical approval was granted. The study was conducted following the STARD 2015 guideline for diagnostic accuracy studies.^30^

Adults presenting to primary care with abdominal symptoms that merited an urgent referral to the NG12 CRC pathway were eligible. People who were under 16 years of age or were unable to understand instructions (including non-English speakers who did not have an interpreter) were not invited to participate.

A FIT kit and a patient information booklet outlining the purpose of the research study were given to patients by a GP, hospital consultant, research nurse or clinical nurse specialist (CNS) where the person was referred for investigation. The patient was asked to take a single sample at their next bowel movement, before completing bowel preparation for colonoscopy or other examination, and post it to a central laboratory. By returning the FIT kit, the patient provided implied consent to participate in the study. Participation did not affect the patients’ clinical care, and they were aware that the FIT result was for research purposes only and they would not be informed of the result.

The FIT kit included (a) a FIT sample collection device in a sealable plastic pouch (OC-Sensor^™^; Eiken Chemical Company, Tokyo, Japan) pre-labelled with the patient’s name, National Health Service (NHS) number, a unique laboratory number and a space to write the sample date, (b) copy of the urgent referral form or patient data sheet (containing information about the patient and the hospital where the examination took place), (c) a patient experience survey consent form and (d) a pre-labelled return envelope. The urgent referral form contained clinical data such as symptoms, reasons for referral, medical history, and sociodemographic factors.

### Sample analysis

Samples were posted to the Clinical Biochemistry department at Barts Health NHS Trust and stored at 4°C prior to analysis, which took place within one week of receipt. Coefficients of variation were 2.8% at 14 μg/g and 3.0% at 91 μg/g. The lower limit of detection was 4 μg/g. The upper analytical limit was 200 μg/g and samples with a concentration above this were not diluted and re-assayed but reported as ≥200 μg/g. The laboratory is accredited by the UK Accreditation Service to ISO 15189 standards. All test results were performed blinded to patient characteristics and cancer outcomes.

### Outcome definition

Clinical outcomes were collected for all patients who provided a viable sample by requesting copies of examination reports from participating sites every month. CRC and other diagnoses were determined by reviewing colonoscopy, flexible sigmoidoscopy, radiology and histology reports, clinic letters and urgent referral forms provided by the participating sites. All diagnoses were verified by medical members of the central research team including the chief investigator. Other bowel pathology was classified as inflammatory bowel disease/colitis/proctitis, diverticulosis/diverticulitis, haemorrhoids, polyps, adenomas (confirmed histologically), high risk adenomas (BSG guideline 2010,^31^), normal and procedure stopped/incomplete.

### Statistical considerations

We aimed to recruit a minimum of 2200 patients to yield at least 80 CRC cases (assuming 3.5% prevalence), which would have an acceptable error rate around a sensitivity of 89% (i.e. 95%CI width of ±6.8%). Sensitivity, false-positive rate (FPR, or 100 minus specificity), positive predictive value (PPV) and negative predictive value (NPV) were calculated as measures of FIT test performance, with 95% confidence intervals (CI) for the key results. Spearman's rank correlation and the Kruskal-Wallis test were used to assess differences in FIT values according to age, sex and ethnicity. Fisher's exact test was used to assess differences in patient features between those with and without CRC cancer. STATA version 15 was used for all analyses.

If a patient returned more than one sample due to being given a test kit in both in primary and secondary care during the same urgent referral or the patient had been investigated more than once, only the first test result was selected for inclusion in the analysis.

### Patient and Public Involvement

Three patient representatives with prior experience with CRC were involved in the development of the patient information leaflet and the design of the FIT kit handout. The participants were invited to complete a patient experience survey (to be published elsewhere).

## RESULTS

### Kit returns and patient characteristics

Within the funded timeframe of the study, FIT kits were returned from 4676 patients in total (which also allowed for the lower cancer prevalence than the 3.5% initially assumed). Among these, 3596 patients provided a viable sample for f-Hb measurement and the outcome after cancer investigations was reported to the co-ordinating centre; all subsequent analyses are based on these (Figure 1). Patient characteristics were similar between the 3596 patients who were included in the analyses and the 1055 who were excluded because their cancer outcome was unknown by the study team (Supplementary Table B).

**Figure 1.**
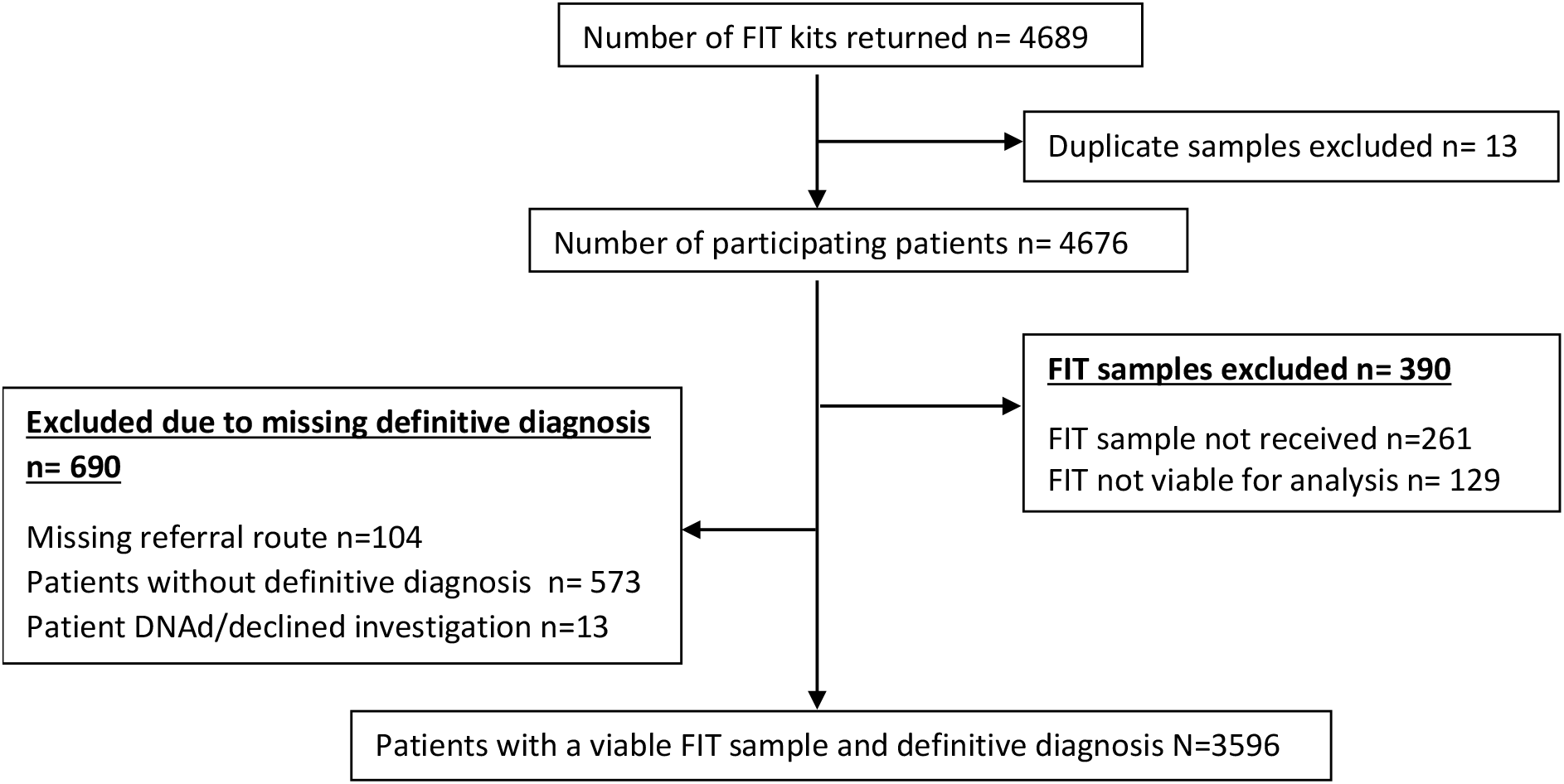
The qFIT study flow diagram

Among the 3596 patients, the majority (99%) were recruited in secondary care. The prevalence of the five most reported clinical features recorded on the urgent referral form within the study cohort were: change of bowel habit 1835 (51%), rectal bleeding 970 (27%), anaemia 684 (19%), abdominal pain 427 (11.9%) and weight loss 312 (8.7%).

The median age was 67 years (70% aged 2265≥60) and 53% were female (Table 1). The association between FIT and each of age, sex and ethnicity were not clinically significant, either in patients with or without CRC (Supplementary Figure A-C). Also, the percentage of people with f-Hb <10 μg/g did not vary according to these three demographic factors.

**Table 1.**
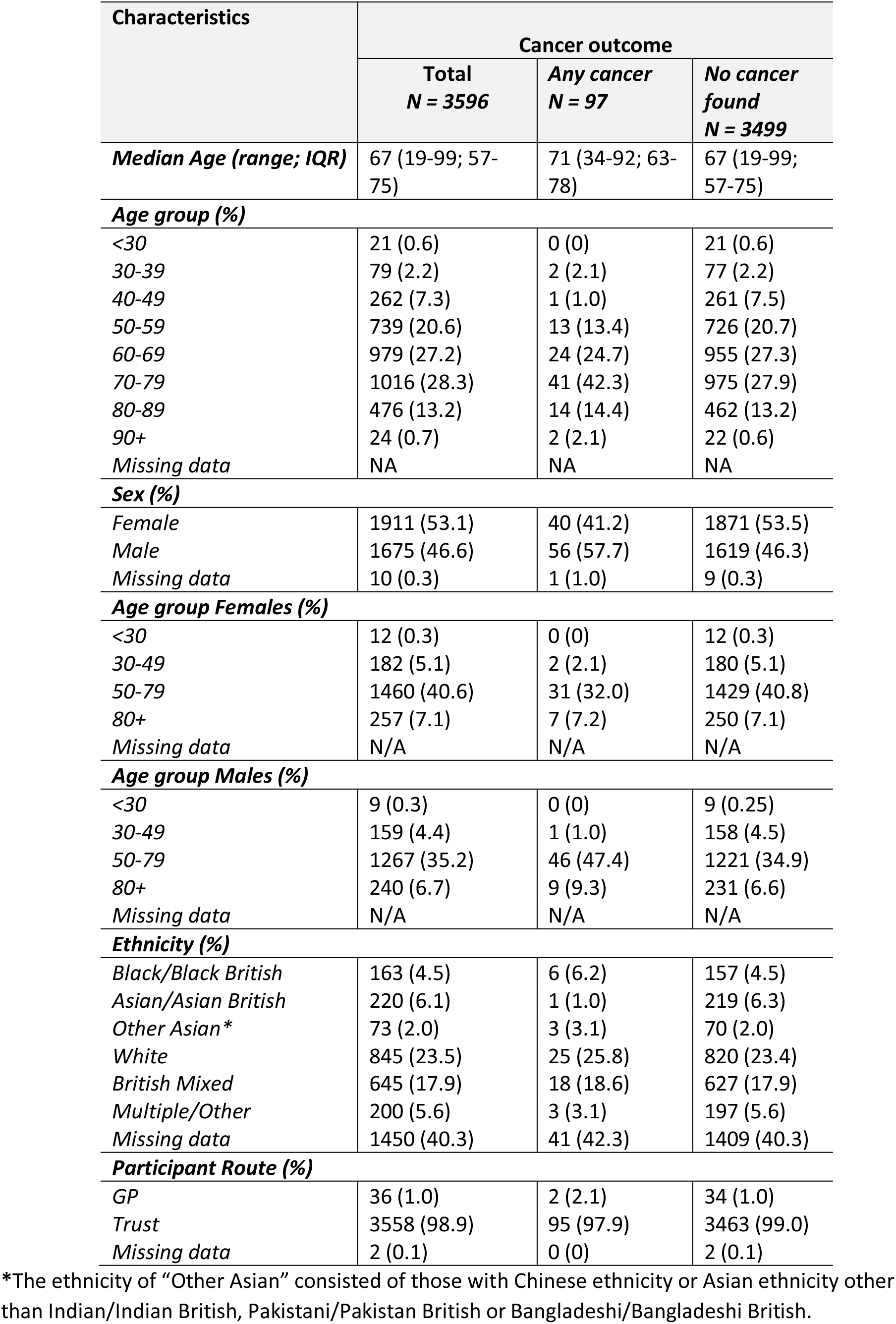
Patient characteristics.

The first investigation recorded was colonoscopy (77.7%), CT colonography (14.2%), flexible sigmoidoscopy (7.5%) (Supplementary Table C). Type of first investigation was related to age; majority of patients over 80 years old were examined using CT colonography, while patients under 80 had a colonoscopy (Supplementary Table C). In those patients undergoing colonoscopy for whom the quality of bowel preparation was recorded, it was satisfactory or better 90.7%.

### Clinical outcomes and FIT test performance

In our study, the NG12 urgent referral pathway had a CRC case finding of 2.5% (90 patients) and all cancers of 2.7% (97 patients), and in 3499 patients (97.3%) no cancer was detected.

The most common diagnoses among patients without cancer were diverticulosis 1101 (31.5%), all polyps 805 (23%), all adenomas 623 (17.8%), high-risk adenomas 61 (1.7%), haemorrhoids 526 (15%), and colitis 286 (8.2%).

Table 2 summarises the performance of the FIT test at different f-Hb cut off values. In our study, the positive predictive value of FIT for CRC ≥10μg/g is almost fourfold that of the NG12 urgent referral pathway (9.7% vs 2.5%). Of the 90 patients with CRC, 15 (16.7%, 95% CI 9.0-24.4) had f-Hb <10 μg/g; hence 1 in 6 cancers would have been missed using this threshold. The characteristics of these 15 patients and their f-Hb values are listed in Table 3. By reducing the f-Hb threshold to 4 μg/g, which was the lowest limit of detection, 12.2% (95%CI 5.5-19.0) of cancers would be missed (1 in 8 cases).

**Table 2.** Test performance of FIT for colorectal cancer (CRC) at different f-Hb cut offs

| Individuals with negative test results, i.e. below the specified f-Hb cut-off |  |  |  |  |  | Individuals with positive test results, i.e. at or above the specified f-Hb cut-off |  |  |  |
| --- | --- | --- | --- | --- | --- | --- | --- | --- | --- |
| FIT Test Result f-Hb cut off (µg/g) | Not Cancer* (n1=3499)<br>True Negatives (TN) | Colorectal Cancer* (n2=90)<br>False Negatives (FN) | Negative Predictive Value (%) (TN/TN+FN) | Risk of CRC among test negatives, (FN/TN+FN) | % of all patients beneath threshold** (N=3596) | FIT Test Result f-Hb cut off (µg/g) | Sensitivity (n2= 90 Colorectal cancers)*<br>True Positives (TP) | False-positive rate* (n1=3499)<br>True Negatives (FP) | Positive Predictive Value (%) (TP/TP+FP) |
|  | No. (%) | No. (%) | (%) | No. per 1000 | (%) |  | % (No.) | % (No.) | (%) |
| <4 | 2556 (73.0) | 11 (12.2) | 99.6 | 4.3 | 71.4 | ≥4 | 87.8 (79) | 27.0 (943) | 7.7 |
| <6 | 2662 (76.1) | 12 (13.3) | 99.6 | 4.5 | 74.4 | ≥6 | 86.7 (78) | 23.9 (837) | 8.5 |
| <10 | 2803 (80.1) | 15 (16.7) | 99.5 | 5.3 | 78.5 | ≥10 | 83.3 (75) | 19.9 (696) | 9.7 |
| <20 | 2993 (85.5) | 17 (18.9) | 99.4 | 5.6 | 83.8 | ≥20 | 81.1 (73) | 14.5 (506) | 12.6 |
| <50 | 3205 (91.6) | 23 (25.6) | 99.3 | 7.1 | 89.9 | ≥50 | 74.4 (67) | 8.4 (294) | 18.6 |
| <80 | 3265 (93.3) | 29 (32.2) | 99.1 | 8.8 | 91.7 | ≥80 | 67.8 (61) | 6.7 (234) | 20.7 |
| <100 | 3294 (94.1) | 32 (35.6) | 99.0 | 9.6 | 92.6 | ≥100 | 64.4 (58) | 5.9 (205) | 22.1 |
| <120 | 3315 (94.7) | 35 (38.9) | 99.0 | 10.4 | 93.3 | ≥120 | 61.1 (55) | 5.3 (184) | 23.0 |
| <150 | 3331 (95.2) | 38 (42.2) | 98.9 | 11.3 | 93.8 | ≥150 | 57.8 (52) | 4.8 (168) | 23.6 |
| <200 | 3347 (95.7) | 41 (45.6) | 98.8 | 12.1 | 94.4 | ≥200 | 54.4 (49) | 4.3 (152) | 24.4 |

\*excludes 7 patients with cancer other than CRC
\*\*90 CRC cases, 7 other cancers and the 3499 without cancer.
False-positive rate is the same as 100 minus specificity

**Table 3.** Primary presenting clinical features and location of tumour in the 15 patients diagnosed with colorectal adenocarcinoma who had f-Hb <10 μg/g (false-negatives)

| No | Sex | Age | f-Hb (µg/g) | Clinical features | Number of clinical features | Location of tumour |
| --- | --- | --- | --- | --- | --- | --- |
| 1 | Male | 60 | <4 | Change in bowel habit;<br>Previous bowel cancer;<br>Rectal bleeding | 3 | Rectum |
| 2 | Female | 70 | <4 | Anaemia; Change in bowel habit | 2 | Pan colonic |
| 3 | Female | 75 | <4 | Abdominal pain; Anaemia;<br>Change in bowel habit | 3 | Sigmoid colon |
| 4 | Male | 67 | <4 | Rectal Bleeding | 1 | Not stated |
| 5 | Male | 92 | <4 | Anaemia; Change in bowel habit | 2 | Descending colon |
| 6 | Female | 53 | <4 | Not stated | Not stated | Not stated |
| 7 | Male | 79 | <4 | Anaemia | 1 | Not stated |
| 8 | Female | 62 | <4 | Abdominal pain; Abnormal imaging; Rectal Bleeding | 3 | Sigmoid colon |
| 9 | Male | 82 | <4 | Anaemia; Rectal Bleeding | 2 | Rectum |
| 10 | Female | 75 | <4 | Anaemia | 1 | Caecum |
| 11 | Male | 74 | <4 | Anaemia; Change in bowel habit | 2 | Not stated |
| 12 | Female | 48 | 4 | Abdominal pain; Change in bowel habit; Family history of CRC | 3 | Splenic flexure |
| 13 | Female | 70 | 7 | Abdominal pain; Change in bowel habit | 2 | Not stated |
| 14 | Male | 64 | 7 | Abdominal pain; Anaemia;<br>Change in bowel habit | 3 | Hepatic flexure |
| 15 | Female | 62 | 7 | Abdominal pain; Change in bowel habit; Weight loss | 3 | Ascending colon |

Using FIT to detect cancer, f-Hb ≥10 μg/g has a sensitivity of 83.3% (95%CI 75.6-91.0), FPR 19.9% (95%CI 18.6-21.1), and PPV of 9.7% (95%CI 7.6-11.8) which represents a 1 in 10 risk of having CRC. Lowering the threshold to ≥4 μg/g modestly increases the sensitivity to 87.8% (95%CI 81.0-94.5) but the FPR increases by 7.1 percentage points to 27.0% (95%CI 25.5-28.4); hence a lower PPV of 7.7% (95%CI 6.1-9.4), or 1 in 13 risk.

A relatively low FPR could be achieved at high thresholds, and at ≥150 μg/g it is 4.8% (95%CI 4.1-5.5), but the sensitivity is 57.8% (95%CI 47.6-68.0). There is no marked increase in CRC risk between f-Hb ≥120 and ≥150 μg/g, with PPVs of 22.1 and 23.6% respectively.

Among 7 patients with other cancers: 2/7, 2/7 and 4/7 had f-Hb <4, <6 and <10 μg/g respectively (Supplementary Table D).

We explored whether the number of missed CRC cases at the lowest FIT thresholds could be minimised by considering patient symptoms and features (listed on the urgent referral form). The percentage of patients with rectal bleeding or change in bowel habit did not discriminate patients with and without CRC. However among all patients with f-Hb<10 μg/g, abdominal pain or anaemia were more likely to be present in cancer patients (66.7%) than in non-cancer patients (29.4%); Table 4. Of the 15 CRC cases missed using f-Hb 10 μg/g alone, only 5 would have been missed in the absence of anaemia or abdominal pain (1 in 18 of all CRC cases; 5.5% [5/90] with 95%CI 0.8-10.3). But at this threshold, 1978 patients without cancer (2803-825; Table 4) did not have these two features, indicating unnecessary further investigations such as colonoscopy could be avoided in 56.5% (1978/3499).

**Table 4.** Distribution of clinical features among individuals with and without CRC at <10 μg/g (False negatives).

| Clinical features | With CRC<br>N=15 (%) | Without CRC<br>N=2803 (%) | p-Value* |
| --- | --- | --- | --- |
| Anaemia | 8 (53.3) | 502 (17.9) | 0.002 |
| Abdominal pain | 6 (40.0) | 355 (12.7) | 0.008 |
| Abdominal pain <u>or</u> anaemia | 10 (66.7) | 825 (29.4) | 0.003 |
| Abdominal pain <u>and</u> anaemia | 2 (13.3) | 16 (0.6) | 0.004 |
| Change in bowel habit | 9 (60.0) | 1506 (53.7) | 0.80 |
| Rectal bleeding | 4 (26.7) | 691 (24.7) | 0.77 |
| Weight loss | 1 (6.7) | 246 (8.8) | >0.99 |
| Family history of CRC | 1 (6.7) | 66 (2.4) | 0.30 |
| Abnormal imaging | 1 (6.7) | 33 (1.2) | 0.17 |
| Previous bowel cancer | 1 (6.7) | 0 (0) | 0.005 |

\*Fisher's exact test

Patients with CRC and low FIT concentrations were also more likely to have multiple symptoms than non-cancer patients (Table 5). 6 of the 15 patients (40%) with CRC who had f-Hb <10 μg/g presented with 3 symptoms or more, while the majority of patients without CRC had none or only one primary symptom (68.7% [1926/2803).

**Table 5.** Comparison of the number of primary clinical features (listed in Table 4) in patients with and without colorectal cancer who had f-Hb <10µg/g

| Number of clinical features | With Colorectal cancer<br>N=15 (#; %) | Without Colorectal cancer<br>N=2803 (#;%) |
| --- | --- | --- |
| 0 | 1 (6.7) | 108 (3.85) |
| 1 | 3 (20.0) | 1818 (64.9) |
| 2 | 5 (33.3) | 686 (24.5) |
| 3 | 6 (40.0) | 191 (6.8) |

Fisher's Exact test p<0.001

## DISCUSSION

This study evaluates use of the quantitative FIT test in people with high-risk symptoms referred on the NG12 urgent suspected CRC pathway, using more patients than any previous study and more CRC cases. The central hypothesis was to examine the ability of FIT, a simple, non-invasive test, to act as a ‘rule out’ tool at the point of referral in primary care, to allow clinicians to triage people with colorectal symptoms into a high risk group warranting urgent investigation, and a lower risk group that could be reassured and managed in the community. Our large study demonstrated that 17% of people with CRC had undetectable or very low levels of faecal haemoglobin (at 10 μg/g cut off level), a proportion higher than in previous, smaller studies, and which we considered to be unacceptable.^18,19,22^ We also showed that FIT levels were not associated with age, sex or ethnicity; unlike some studies of asymptomatic people.

NPV is often reported in the literature as a measure of test performance. However, very high NPVs can be due to having a large number of non-cancer patients in relation to a small number of CRC cases, particularly in small studies. The NPV in our study was 99.5% at a FIT threshold of <10 μg/g, similar to other studies^18–21,32^ but this masks the finding that as many as 17% of cancers would have been missed. High NPVs may therefore give false reassurance about the effectiveness of the FIT test in ruling out CRC.

Our study is important because it is a real world assessment of FIT as a triage tool for primary care. Participants were drawn from a wide geography representing a diverse demographic across primary and secondary care. The cancer investigations were representative of pragmatic clinical practice; for example, the increased use of CT colonography for those over 80 years of age.

Although 17% of CRC detected by the NG12 pathway would not have been identified by FIT alone, 7 of the 11 patients with cancer with f-Hb <4 μg/g had anaemia as the presenting symptom, and 8 of the 15 patients with cancer with f-Hb <10 μg/g had anaemia as the presenting symptom (Table 3). Our results suggest that clinicians should be encouraged to safety net a negative FIT result with clinical features such as anaemia and abdominal pain. Small prospective studies^20,33^ and cohort studies^10,34^ have similarly identified the association of anaemia with FIT when identifying patients with CRC.

Interestingly, rectal bleeding and change in bowel habit were not significant discriminators for patients with or without CRC and FIT <10μg/g. In addition to anaemia, abdominal pain was the second most significant clinical feature in CRC patients who had low f-Hb (Table 4). Using both of these clinical features in addition to FIT, the missed cancer rate at f-Hb <10 μg/g reduced from 17% to 5.5%. These figures should be compared with the reported three-year post-colonoscopy CRC rate in England which was 7.4%.^35^ A novel outcome from our study was that the number of clinical features also appear predictive of colorectal cancer at f-Hb <10 μg/g (Table 5). Consideration of these findings should help inform an evidence-based approach to safety netting, to minimise the number of CRC cancers missed by FIT if this test were to be used in routine practice in high risk patients.

Our study had a few limitations. 2.9% of our samples were unsuitable for FIT analysis. This is in line with previous studies that demonstrate issues over stool self-sampling, such as delay in posting the sample back^36^ or picking the stool.^37^ Only a single stool sample was requested from each patient, and Hogberg et al (2017) indicate that this could lead to missing one tenth of symptomatic CRCs and adenomas with high grade dysplasia^38^, compared to using three samples; while other studies did not find any significant improvement in test accuracy when two FITs were performed.^21,39^ A final clinical diagnosis could not be obtained from records on all patients during the study period. As a consequence, several patients with a valid FIT result were excluded from the final analysis. However, the characteristics of these patients were similar to those included in the analyses (Supplementary Table B) so selection bias is unlikely to have occurred. FIT results could be affected by genetic disorders including haemoglobinopathy and Lynch syndrome, and medications such as aspirin, iron, non-steroidal anti-inflammatory drugs or anticoagulants. These data were not sufficiently available for full analysis. Finally, it would have been useful to examine cancer stage, particularly in those who had low FIT values to see whether most were early stage, but this information was unavailable at the time of analysis.

This study used the OC-Sensor™iO with a lower limit of detection of 4 μg/g, which was one of the recommended analysers in the NICE DG30 guidance.^23^ NICE suggests that the three recommended analysers are comparable when used in line with its DG30 guidance. However, it is known that there are differences in analytical performance^40^ which may affect generalisability of results between studies. The relevance of this is unknown, but it is a potential limitation. The relevance of the lower limit of detection is speculative but it would be reasonable to assume that reducing this would improve the sensitivity of FIT, at the expense of increasing false positives.

Employing FIT as a triage tool can target potentially unpleasant and harmful investigations for patients who would most benefit; f-Hb thresholds of ≥10 μg/g and ≥4 μg/g, could potentially allow 80% and 73% patients to avoid further investigations, respectively, which is similar to previous findings.^24,39^ However, the missed CRC rate is too high using FIT on its own, but can be minimised by referring patients with anaemia or abdominal pain despite having a low FIT result, thus ensuring safety netting using these clinical features. This approach could result in only 44% of symptomatic patients without cancer being investigated further (using our findings). This represents a significant saving in healthcare resources, and would address the concerns that NG12 increases referrals and overburdens endoscopy and radiology departments without substantially improving the CRC diagnostic rate.^41^

Practical clinical use of FIT as a triage tool will clearly be hampered by concerns of both clinicians and patients about missed cancers (false-negative FIT). In the absence of a validated clinical risk score a viable strategy for safety netting would be a second clinical review to allow reassessment of the patient. Although this study does not provide definitive guidance of how this reassessment should be conducted, the fact that the presence of multiple clinical features, particularly anaemia or abdominal pain, appear to be more common in FIT negative cancers would suggest a repeat blood count. Demonstrating a drop in haemoglobin, alongside additional clinical features, could be used to consider the patient for urgent referral. Further studies should evaluate the value of repeat FIT if symptoms persist as part of a safety netting option.

To conclude, this study demonstrates that FIT is a powerful triage tool with a low false-positive rate and high sensitivity for patients presenting with high risk symptoms of CRC. However, FIT alone is an imperfect test to rule out colorectal cancer in patients presenting with high risk colorectal symptoms in primary care. Provision of an adequate, evidence-based safety netting system is required to identify FIT-negative cancers. Importantly, the utility of FIT is increased by consideration of clinical features, and as such it has the ability to focus secondary care diagnostic investigations on the patients with the most need and potentially delivering a CRC miss rate lower than that of colonoscopy.

#### in the box

What is already known on this topic
- NICE recommends patients with ‘high risk’ lower gastrointestinal symptoms are referred onto an urgent pathway for further investigation, with an expected CRC rate of 3%
- The faecal immunochemical test (FIT) is used in England for screening and to triage patients with ‘low risk’ bowel symptoms for investigation to diagnose colorectal cancer (CRC).
- To date, there is insufficient evidence for NICE to recommend using FIT to triage ‘high risk’ symptomatic patients on the urgent CRC pathway. Previous studies were too small to reliably estimate the number of missed cancers.

What this study adds
- We conducted one of the largest ever studies to evaluate FIT (including 90 CRC cases).
- The positive predictive value of FIT for CRC ≥10μg/g is almost fourfold that of the NG12 urgent referral pathway (9.7% vs 2.5%).
- But FIT alone at f-Hb 10μg/g would mean that 17% of CRC cancers are missed.
- Clinical features assist safety netting: using FIT 10μg/g in conjunction with anaemia and abdominal pain would lead to only 5.5% of CRC cancers being missed.
- Rectal bleeding and change in bowel habit were not significant discriminators for patients with or without CRC who had FIT <10μg/g.

## POLICY IMPLICATIONS AND CONCLUSIONS

- FIT with clinical features is a powerful triage tool with high specificity and sensitivity for patients presenting with high risk symptoms of CRC.
- If selection for urgent investigation among high risk patients had been based on FIT alone at f-Hb ≥10 μg/g, 80% lower abdominal investigations could have been avoided in patients who do not have cancer, but 17% of CRC cases would have been missed.
- If FIT is to be commissioned as a triage tool in high risk patients, safety netting with clinical features must be part of the urgent referral pathway. By considering anaemia and abdominal pain among patients who have f-Hb <10 μg/g, only 5% of CRC would be missed and 56% of people without cancer could avoid a lower abdominal investigation.
- Implementation studies can confirm the utility of combining FIT with clinical features for safety netting, the value of repeat FIT testing, and to quantify the reduction in healthcare resource use.

## CONTRIBUTORS

HEL formulated the study design and protocol, managed the study, drafted and finalised the manuscript. ES, RA and KPJ advised on to the study protocol. MM, ES, DC and KPJ conducted medical review of investigations. RA and JL conducted the FIT test analysis. MM, ES, CS, RA, AM, AH, DC and KPJ contributed to the interpretation of results and the critical revision of the manuscript. AM and AH conducted the statistical analysis. CS and MM contributed to the recruitment of participants. DC advised on the data management and conducted data cleaning. MM was chief investigator of the study, initiated the study concept and provided oversight and is the guarantor.

## Data Availability

Additional summary tables and analyses are available upon request from the senior author, Mr Michael Machesney.

## ACKNOWLEDGEMENT

The authors would like to thank all participating sites for allocating resources to carry out patient recruitment and follow up, former data managers Safreena Nazir and James Croft for their data extraction work, Sara Taiyari for recruiting the most participants at multiple sites, Jennifer McGivney, Ed Carbonell, Marcy Madzikanda, Mayada Kaddoura, Dimple Makwana and Hasmita Shivji for the administrative support during the conducting of the study, Aska Matsunaga at NOCLOR for her support in recruiting GP practices and Claire Levermore, Mairead Lyons and Nick Kirby for encouragement and allocation of resources to perform the study as part of the National Cancer Vanguard New Care Models programme.

## DISCLAIMER

The funding sources had no role in the design, conduct or reporting of the study.

## ETHICS APPROVAL

Ethical approval was granted by NRES West Midlands - Solihull Research Ethics Committee (Ref. 17/WM/0094) and the Health Research Authority (IRAS/213710).

## PATIENT CONSENT FOR PUBLICATION

Not required

## DATA SHARING STATEMENT

Additional summary tables and analyses are available upon request from the corresponding author.

## Supplementary figures and tables

### Supplementary Figures

**Figure A.**
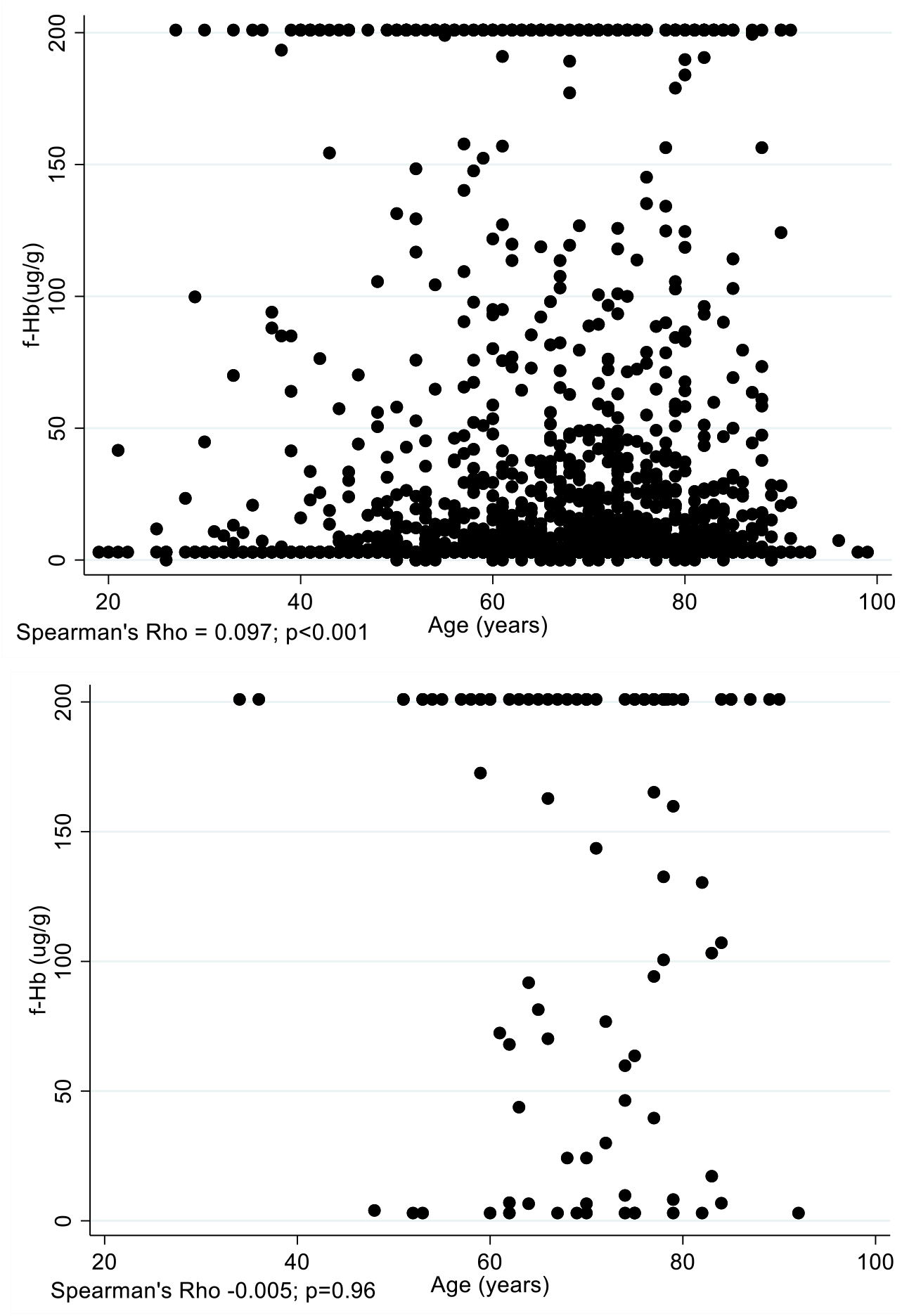
Association between the FIT test result and age among 3499 patients without cancer (upper figure) and 97 with cancer (lower figure). Due to the large sample size of non-cancers, the clinically insignificant correlation of 0.097 is statistically significant.

**Figure B.**
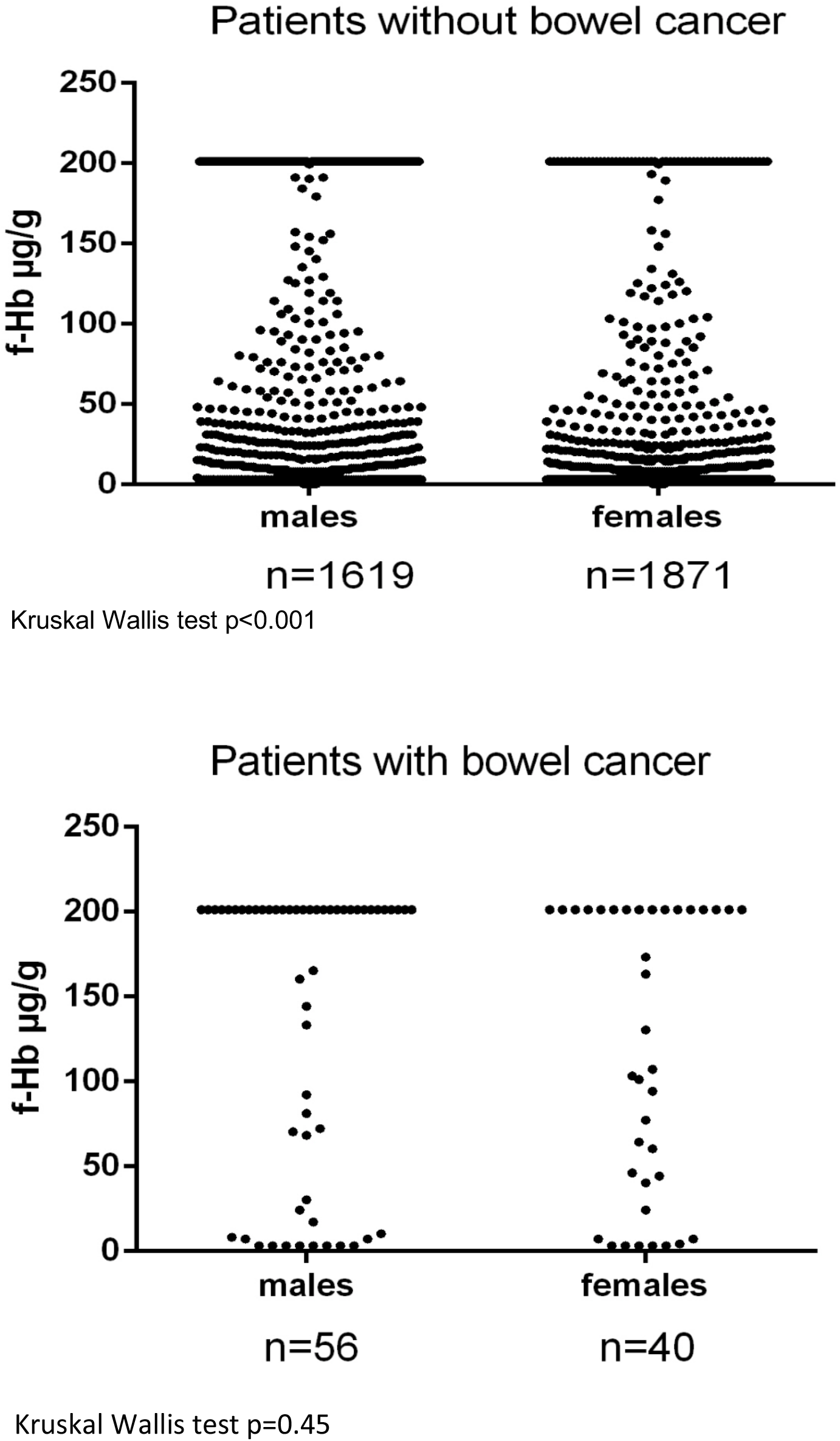
Association between the FIT test result and sex among 3490 patients without cancer (upper figure) and 97 with cancer (lower figure). In those without cancer the median f-Hb is <4 ug/g (males) and <4 ug/g (females). Due to the large sample size of non-cancers, a clinically insignificant difference is statistically significant. In those with cancer the median f-Hb is >200 ug/g (males) and 146.5 ug/g (females). Among all patients, 74.8% of males had f-Hb<10 μg/g, compared to 81.7% of females (p<0.001), but this risk ratio 0.92 (74.8/81.7) is not considered clinically significant.

**Figure C.**
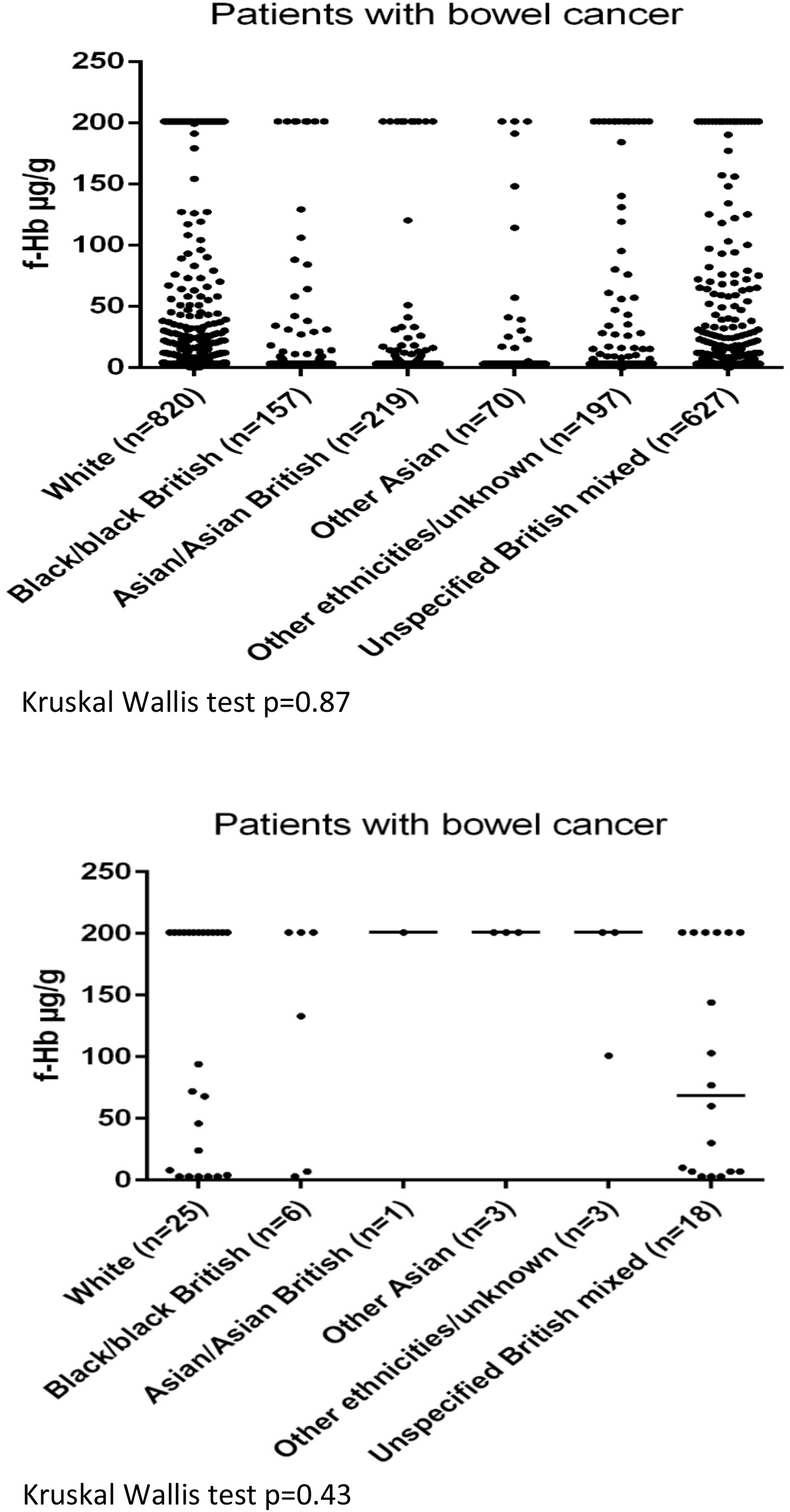
Association between the FIT test result and ethnicity among 2090 patients without cancer (upper figure) and 56 with cancer (lower figure). Among all patients, the percentage that had f-Hb<10 μg/g was 31.0% white, 6.2% black, 8.8% Asian, 2.6% other Asian, 7.3% Other ethnicities/unknown and 23.5% unspecified British mixed (p=0.28).

### Supplementary tables

**Table A.**
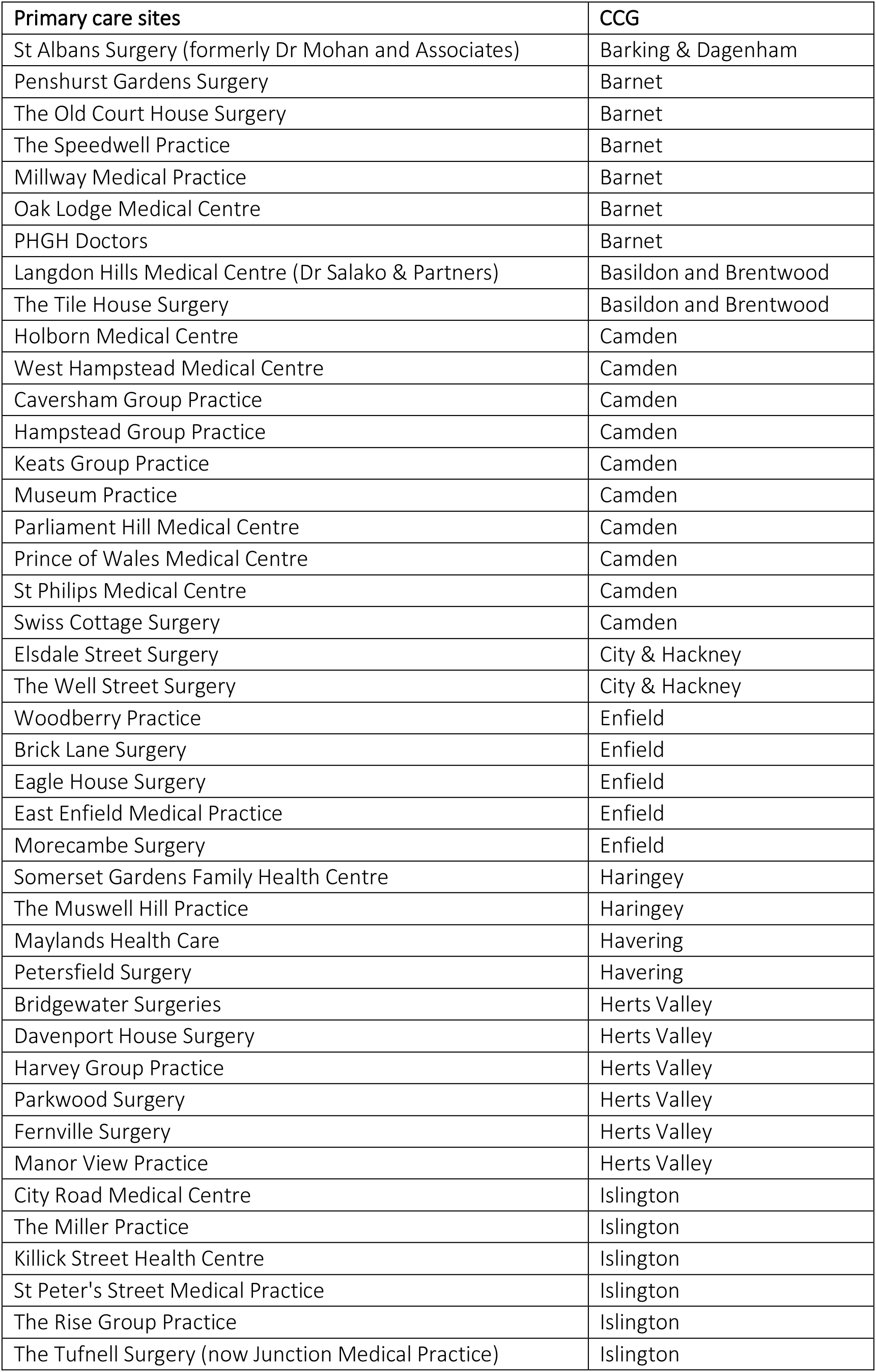
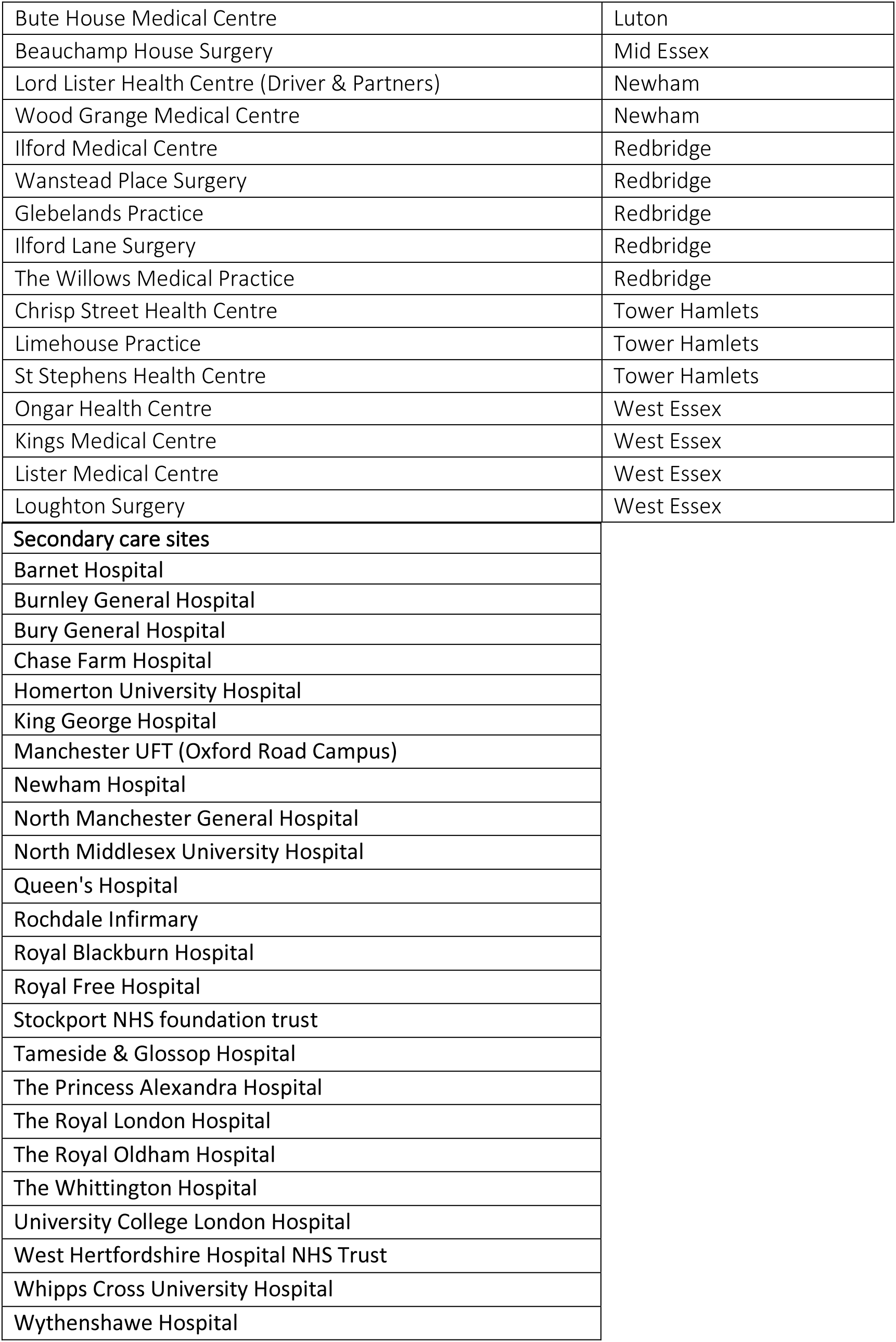
List of participating sites in the qFIT study

**Table B.** A comparison of the characteristics in patients that had a definitive diagnosis and a viable FIT sample, with patients where the definitive diagnosis was unknown by the study team

| **Characteristics** | ***Known definitive diagnosis (used in analysis) N=3596*** | ***Missing definitive diagnosis N=1055*** |
| --- | --- | --- |
| ***Median Age (range; IQR)*** | 67 (19-99; 57-75) | 70 (17-100; 58-78) |
| ***Median f-HB (N); (range; IQR)*** | <4 (N=3596)<br>(<4 - >200; <4 - 6) | <4 (N=696)<br>(<4 - >200; <4 - 7) |
| *qFIT value <10 µg/g (%)* | 2852 (79.3) | 549 (78.9) |
| *qFIT value <20 µg/g (%)* | 3026 (84.1) | 593 (85.2) |
| ***Age group (%)*** |  |  |
| <30 | 21 (0.6) | 11 (1.1) |
| 30-39 | 79 (2.2) | 24 (2.4) |
| 40-49 | 262 (7.3) | 72 (7.2) |
| 50-59 | 739 (20.6) | 165 (16.4) |
| 60-69 | 979 (27.2) | 232 (23.1) |
| 70-79 | 1016 (28.3) | 298 (29.7) |
| 80-89 | 476 (13.2) | 188 (18.7) |
| 90+ | 24 (0.7) | 15 (1.5) |
| ***Sex (%)*** |  |  |
| *Female* | 1911 (53.3) | 430 (47.9) |
| *Male* | 1675 (46.7) | 468 (52.1) |
| ***Ethnicity (%)*** |  |  |
| *Black/Black British* | 163 (7.6) | 23 (6.6) |
| *Asian/Asian British* | 220 (10.3) | 37 (10.5) |
| *Other Asian\** | 73 (3.4) | 13 (3.7) |
| *White* | 845 (39.4) | 153 (43.6) |
| *British Mixed* | 645 (30.1) | 98 (27.9) |
| *Multiple/Other* | 200 (9.3) | 27 (7.7) |

\*The ethnicity of "Other Asian" consisted of those with Chinese ethnicity or Asian ethnicity other than Indian/Indian British, Pakistani/Pakistan British or Bangladeshi/Bangladeshi British.
Missing values for characteristics were not included in the table.

**Table C.** First type of exam by age group (in patients with a definitive diagnosis and a valid FIT test) N=3585

| Type of exam | Age group | Number of patients that had exam (N; %) | Total N=3596 |
| --- | --- | --- | --- |
| Colonoscopy | <30<br>30-49<br>50-79<br>80+ | 17 (0.6)<br>300 (10.7)<br>2239 (80.1)<br>238 (8.5) | 2794 |
| CT Colonography | <30<br>30-49<br>50-79<br>80+ | 0 (0)<br>11 (2.2)<br>312 (61.1)<br>188 (36.8) | 511 |
| Flexible Sigmoidoscopy | <30<br>30-49<br>50-79<br>80+ | 4 (1.5)<br>29 (10.8)<br>168 (62.7)<br>67 (25.0) | 268 |
| CT Scan | <30<br>30-49<br>50-79<br>80+ | 0 (0)<br>0 (0)<br>3 (60.0)<br>2 (40.0) | 5 |
| Rigid Sigmoidoscopy | <30<br>30-49<br>50-79<br>80+ | 0 (0)<br>0 (0)<br>1 (100)<br>0 (0) | 1 |
| Other | <30<br>30-49<br>50-79<br>80+ | 0 (0)<br>1 (25.0)<br>2 (50.0)<br>1 (25.0) | 4 |
| Not Applicable | <30<br>30-49<br>50-79<br>80+ | 0 (0)<br>0 (0)<br>1 (50.0)<br>1 (50.0) | 2 |
| Missing Data | <30<br>30-49<br>50-79<br>80+ | 0 (0)<br>0 (0)<br>8 (72.7)<br>3 (27.3) | 11 |

**Table D.**
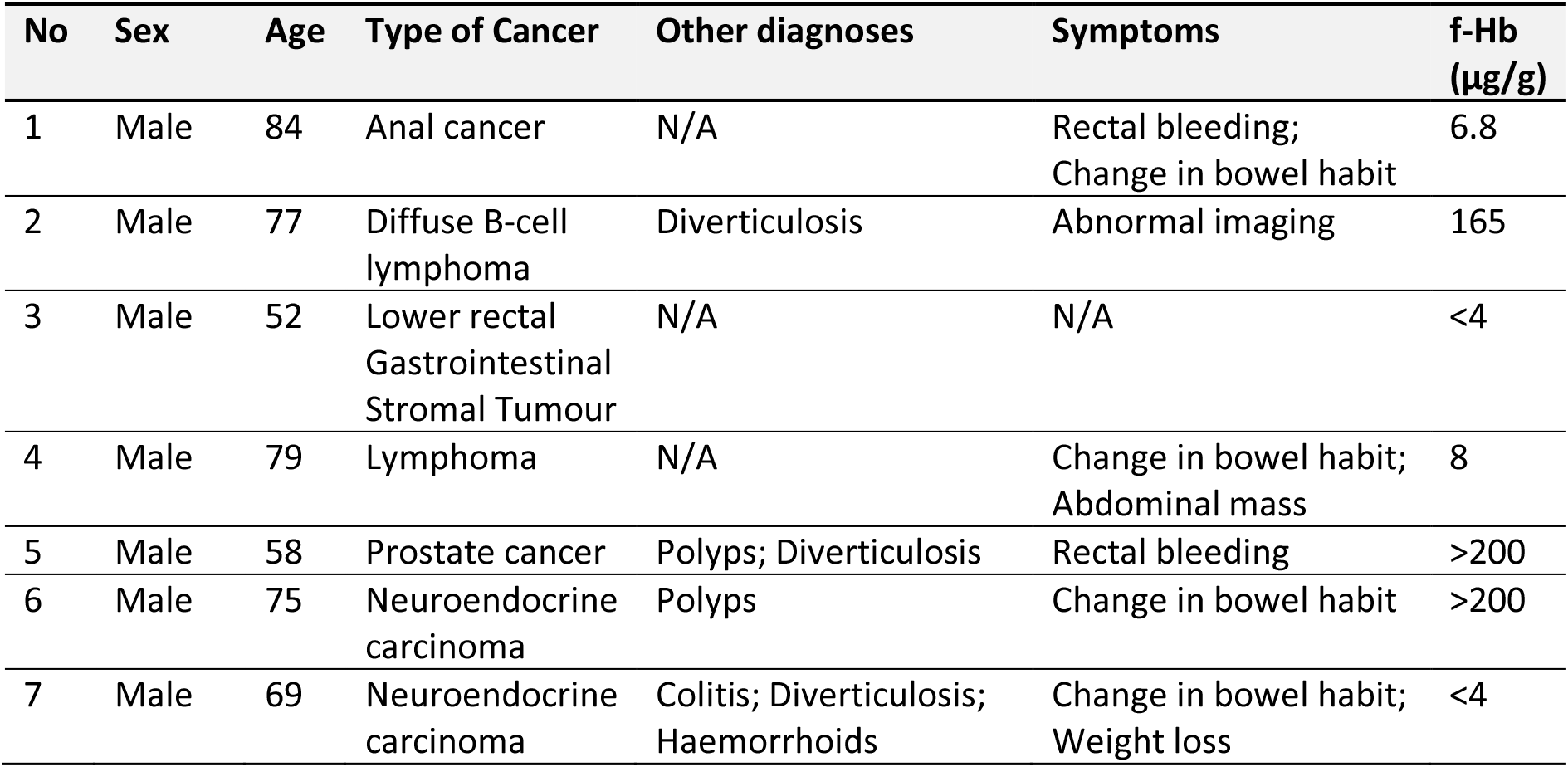
Patients found to have cancers other than colorectal adenocarcinoma and their respective other diagnoses, symptoms and FIT concentrations (μg/g)

| No | Sex | Age | Type of Cancer | Other diagnoses | Symptoms | f-Hb (µg/g) |
| --- | --- | --- | --- | --- | --- | --- |
| 1 | Male | 84 | Anal cancer | N/A | Rectal bleeding;<br>Change in bowel habit | 6.8 |
| 2 | Male | 77 | Diffuse B-cell lymphoma | Diverticulosis | Abnormal imaging | 165 |
| 3 | Male | 52 | Lower rectal Gastrointestinal Stromal Tumour | N/A | N/A | <4 |
| 4 | Male | 79 | Lymphoma | N/A | Change in bowel habit;<br>Abdominal mass | 8 |
| 5 | Male | 58 | Prostate cancer | Polyps; Diverticulosis | Rectal bleeding | >200 |
| 6 | Male | 75 | Neuroendocrine carcinoma | Polyps | Change in bowel habit | >200 |
| 7 | Male | 69 | Neuroendocrine carcinoma | Colitis; Diverticulosis; Haemorrhoids | Change in bowel habit;<br>Weight loss | <4 |

